# Factors Associated with the Frequency of Medical Consultations in 1,355,354 Patients with Various Types of Diabetes Mellitus

**DOI:** 10.1101/2024.06.15.24308977

**Authors:** Víctor Juan Vera-Ponce, Joan A. Loayza-Castro, Rafael Tapia-Limonchi, Enrique Vigil-Ventura

## Abstract

**Introduction:** Proper management of diabetes mellitus (DM) is essential to prevent long-term complications, improve patients’ quality of life, and reduce the economic burden on healthcare services.

**Objective:** To determine the factors associated with the number of medical consultations received in the last quarter by patients with different types of diabetes affiliated with the Comprehensive Health Insurance (SIS: acronym in Spanish) in Peru.

**Methods:** A cross-sectional analysis of the database of patients with DM affiliated with SIS in Peru was conducted. Two robust variance regression models were used to identify potential associated factors.

**Results:** Data from 1,355,354 patients were analyzed. In model 1, which included comorbidities as separate variables, it was found that men, older individuals (especially those aged 60-69), and residents of the jungle region had a higher probability of receiving more medical consultations. The presence of obesity/dyslipidemia, hypertension, and mental health disorders also increased the likelihood of more consultations. In comparison, patients with type 2 DM had fewer consultations compared to those with type 1 DM. The findings were consistent with the first model in model 2, which included the total number of comorbidities instead of each separately. Additionally, a higher total number of comorbidities was associated with an increased number of medical consultations.

**Conclusions:** Several vital factors influencing the frequency of medical consultations received by DM patients have been identified. Adapting healthcare services to address regional disparities in access to and use of medical services is crucial.

## Introduction

Diabetes mellitus (DM) is a chronic disease that affects millions of people worldwide and poses a significant challenge to healthcare systems ^(1)^. Proper diabetes management is essential to prevent long-term complications, improve patients’ quality of life, and reduce the economic burden on healthcare services ^(2)^.

Patients with this condition require continuous monitoring and regular medical attention to control their blood glucose levels, manage comorbidities, and prevent acute and chronic complications ^(3)^. However, medical care frequency and quality vary considerably among patients, influenced by clinical, demographic, and socioeconomic factors ^(4–7)^. There is a need to understand whether different aspects such as sex, age, duration of diagnosis, and the presence of comorbidities like obesity and hypertension can influence the frequency of medical consultations. This need is particularly pronounced in different geographical contexts and healthcare systems where the reach of patient care is limited in some areas, as is the case in Peru ^(8)^.

Therefore, the primary objective of this study is to determine the factors associated with the number of medical consultations received in the last quarter by patients with different types of diabetes affiliated with the Comprehensive Health Insurance (SIS: acronym in Spanish) in Peru.

## Methods

### Type and Study Design

This is an analytical cross-sectional study. An open-access database providing detailed information on the demographic and clinical characteristics of patients with DM affiliated with SIS in Peru was analyzed.

### Population, Sample, and Eligibility Criteria

The study sample consisted of patients who met the following selection criteria: 1) patients affiliated with SIS with a definitive diagnosis of DM (type 1, type 2, or other types of diabetes); 2) patients who had received medical consultations in the last three months according to SIS database records.

### Variables and Measurement

The dependent variable in this study is the number of medical consultations received by DM patients in the last quarter. This variable was categorized into two groups: patients who received up to two medical consultations and those who received three or more in the previous quarter.

The independent variables in this study were selected based on their potential influence on the number of medical consultations received by DM patients. The first of these variables is the patient’s sex, classified as male or female. The patient’s age was categorized into groups: under 18 years, and then every 10 years up to 70 years and older. The geographical region was divided into Metropolitan Lima, the rest of the coast, the highlands, and the jungle. The time since diagnosis, referring to the duration since the DM diagnosis, was categorized in yearly intervals up to 6 years and more. Additionally, the presence of comorbidities such as DM with obesity/dyslipidemia, DM with hypertension (HTN), and DM with mental health disorders were included. Finally, the type of DM diagnosed was classified into type 1 (T1DM), type 2 (T2DM), and other specific types of diabetes.

### Procedures

The open-access SIS database of Peru was downloaded from the website https://www.datosabiertos.gob.pe/dataset/afiliados-activos-en-el-seguro-integral-de-salud-con-diagn%C3%B3stico-de-diabetes-mellitus-sis. This database contains nominal information on active affiliates with a definitive diagnosis of diabetes mellitus, including details on medical consultations received in the last three months. The downloaded data were analyzed, and valuable variables for the study were integrated.

Patients registered in the database were evaluated and classified according to their type of DM (type 1, type 2, or other types). Additionally, it was determined if the patients had HTN, and laboratory tests were performed to measure their glucose, lipid levels, and other relevant variables. Patients’ weight and height were measured to calculate their body mass index (BMI) and classify those with obesity (BMI ≥ 30 kg/m2). Mental health was also assessed. These evaluations were conducted in the regions where SIS operates, and the results were stored in the database used for this study.

The study sample included patients affiliated with SIS with a definitive diagnosis of DM who had received at least one medical consultation in the last three months. Patients whose DM diagnosis was not specified as type 1, type 2, or other specific types were excluded to ensure data accuracy. The International Classification of Diseases, Tenth Revision (ICD-10) was used for comorbidity classification. DM with obesity/dyslipidemia was classified under ICD-10 codes E660-E669 and E780-E785. DM with hypertension was classified under ICD-10 codes I10X, I150, I151, I152, I158, and I159. DM with mental health disorders was classified under ICD-10 codes F000-F09X and F100-F259, F300-F99X, respectively.

### Statistical Analysis

An initial descriptive analysis was conducted to characterize the study sample. This analysis included describing demographic and clinical variables such as sex, age, region of residence, time since diabetes diagnosis, presence of comorbidities, and the number of medical consultations received in the last quarter. A bivariate analysis was also performed to explore the relationships between the dependent variable (number of medical consultations received) and each independent variable.

Finally, a Poisson regression analysis with robust variance was conducted to obtain adjusted prevalence ratios (aPRs). Each association measure was accompanied by its respective 95% confidence interval (95% CI) to estimate the precision of the results. Additionally, two distinct models were developed:

- **Model 1 (comorbidities separately)** In this model, comorbidities such as hypertension, obesity, and mental health problems were included as separate variables. This approach allows for identifying the specific impact of each comorbidity on the number of medical consultations received.
- **Model 2 (total number of comorbidities)** This second model included a variable summarizing the total number of comorbidities. This variable represents a cumulative measure of the comorbidity burden, simplifying the model and facilitating the interpretation of the total burden. This approach allows the evaluation of the combined effect of comorbidities rather than their individual effects.

Performing both approaches provides robustness verification. Comparing the results of both models will determine if the conclusions are consistent regardless of the approach used. If both models produce consistent results, the confidence in the findings will be strengthened. Conversely, if the results differ, it may indicate that how comorbidities are modeled significantly affects the results. This difference is crucial for interpretation and will help better understand how the comorbidity burden influences the number of medical consultations received.

### Ethical Aspects

The study adhered to all ethical and privacy regulations. Although an open-access database was used, patient data confidentiality and anonymity were guaranteed. These procedures enabled a thorough and rigorous analysis, providing valuable information to improve the management and care of this disease within the healthcare system.

## Results

The study sample consisted of 1,355,354 patients. The majority of the patients were female (66.13%). Regarding age distribution, the largest group was patients aged 50-59 (24.35%). In terms of diabetes type, an overwhelming majority of patients had T2DM (91.80%). Geographically, Metropolitan Lima housed 37.37% of the total; as for comorbidities, 64.46% of patients had obesity or dyslipidemia, while 38.70% had hypertension. Mental health disorders were less common, affecting 21.78% of patients. Finally, only 9.14% presented all three comorbidities considered in the study when observing the total number of comorbidities.

**Table 1.** Univariate and Bivariate Descriptive Characteristics of the Study Sample.

| Characteristic | Total | Number of medical consultations |  |
| --- | --- | --- | --- |
|  | n = 1,355,354 | < 3<br>n = 1,318,874 | ≥ 3<br>n = 36,480 |
| Sex |  |  |  |
| Female | 896,302 (66.13%) | 871,516 (66.08%) | 24,786 (67.94%) |
| Male | 459,052 (33.87%) | 447,358 (33.92%) | 11,694 (32.06%) |
| **Edad** |  |  |  |
| Under 18 years | 30,146 (2.22%) | 30,010 (2.28%) | 136 (0.37%) |
| 18 - 29 years | 63,267 (4.67%) | 62,913 (4.77%) | 354 (0.97%) |
| 30 - 39 years | 104,003 (7.67%) | 102,552 (7.78%) | 1,451 (3.98%) |
| 40 - 49 years | 218,914 (16.15%) | 214,032 (16.23%) | 4,882 (13.38%) |
| 50 - 59 years | 329,998 (24.35%) | 320,204 (24.28%) | 9,794 (26.85%) |
| 60 - 69 years | 321,288 (23.71%) | 309,964 (23.51%) | 11,324 (31.04%) |
| 70 years and older | 287,494 (21.22%) | 278,956 (21.15%) | 8,538 (23.41%) |
| **Type of Diabetes** |  |  |  |
| Type 1 diabetes | 90,380 (6.67%) | 87,131 (6.61%) | 3,249 (8.91%) |
| Type 2 diabetes | 1,244,198 (91.80%) | 1,211,615 (91.87%) | 32,583 (89.32%) |
| Other specified types of DM | 20,776 (1.53%) | 20,128 (1.53%) | 648 (1.78%) |
| **Region** |  |  |  |
| Metropolitan Lima | 506,526 (37.37%) | 495,520 (37.57%) | 11,006 (30.17%) |
| Rest of the Coast | 496,984 (36.67%) | 485,047 (36.78%) | 11,937 (32.72%) |
| Highlands | 189,034 (13.95%) | 182,011 (13.80%) | 7,023 (19.25%) |
| Jungle | 162,810 (12.01%) | 156,296 (11.85%) | 6,514 (17.86%) |
| **Time since diagnosis** |  |  |  |
| 0 to 1 year | 50,569 (3.73%) | 48,936 (3.71%) | 1,633 (4.48%) |
| 1 to 2 year | 288,794 (21.31%) | 281,428 (21.34%) | 7,366 (20.19%) |
| 2 to 3 year | 239,160 (17.65%) | 234,321 (17.77%) | 4,839 (13.26%) |
| 3 to 4 year | 149,786 (11.05%) | 146,639 (11.12%) | 3,147 (8.63%) |
| 4 to 5 year | 202,075 (14.91%) | 197,731 (14.99%) | 4,344 (11.91%) |
| 5 to 6 year | 243,926 (18.00%) | 237,618 (18.02%) | 6,308 (17.29%) |
| More tan 6 years | 181,044 (13.36%) | 172,201 (13.06%) | 8,843 (24.24%) |
| **DM + Obesity/Dyslipidemia** |  |  |  |
| No | 481,688 (35.54%) | 475,789 (36.08%) | 5,899 (16.17%) |
| Yes | 873,666 (64.46%) | 843,085 (63.92%) | 30,581 (83.83%) |
| **DM + Hypertension** |  |  |  |
| No | 830,800 (61.30%) | 815,672 (61.85%) | 15,128 (41.47%) |
| Yes | 524,554 (38.70%) | 503,202 (38.15%) | 21,352 (58.53%) |
| **DM + Mental Health Disorder** |  |  |  |
| No | 1,060,093 (78.22%) | 1,035,952 (78.55%) | 24,141 (66.18%) |
| Yes | 295,261 (21.78%) | 282,922 (21.45%) | 12,339 (33.82%) |
| **Number of Comorbidities** |  |  |  |
| None | 300,438 (22.17%) | 297,786 (22.58%) | 2,652 (7.27%) |
| One | 540,215 (39.86%) | 529,119 (40.12%) | 11,096 (30.42%) |
| Two | 390,837 (28.84%) | 375,817 (28.50%) | 15,020 (41.17%) |
| Three | 123,864 (9.14%) | 116,152 (8.81%) | 7,712 (21.14%) |

In Model 1, comorbidities were included as separate variables. Men, older individuals (especially those aged 60-69), and residents of the jungle had a higher probability of receiving more consultations. Patients with type 2 DM had a lower probability compared to those with type 1 DM. Additionally, the presence of obesity/dyslipidemia, hypertension, and mental health disorders was significantly associated with an increase in consultations.

Model 2 included a variable representing the total number of comorbidities instead of each comorbidity separately. The findings were consistent with Model 1, and it was also found that a higher total number of comorbidities was strongly associated with increased medical consultations.

**Table 2.**
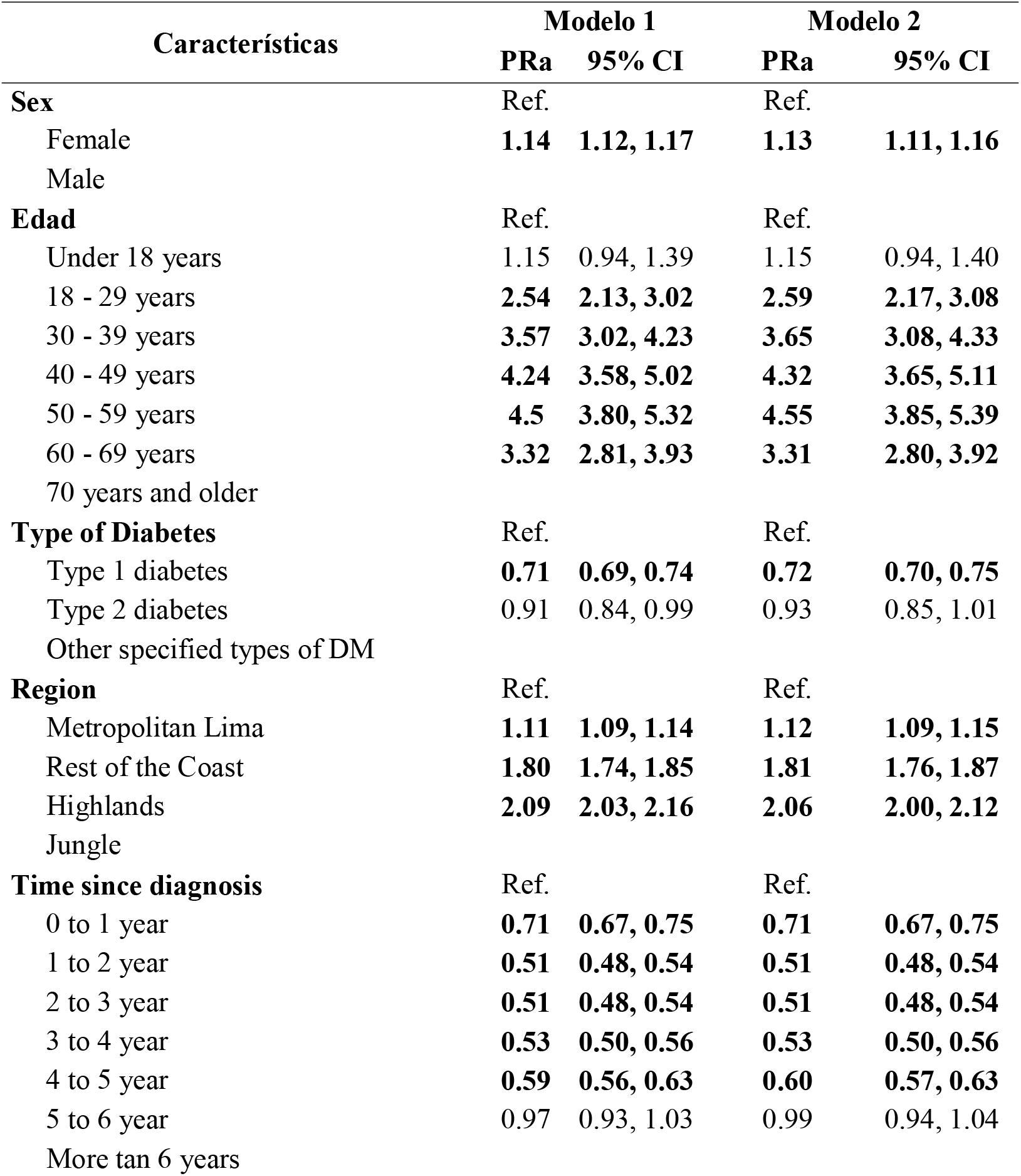
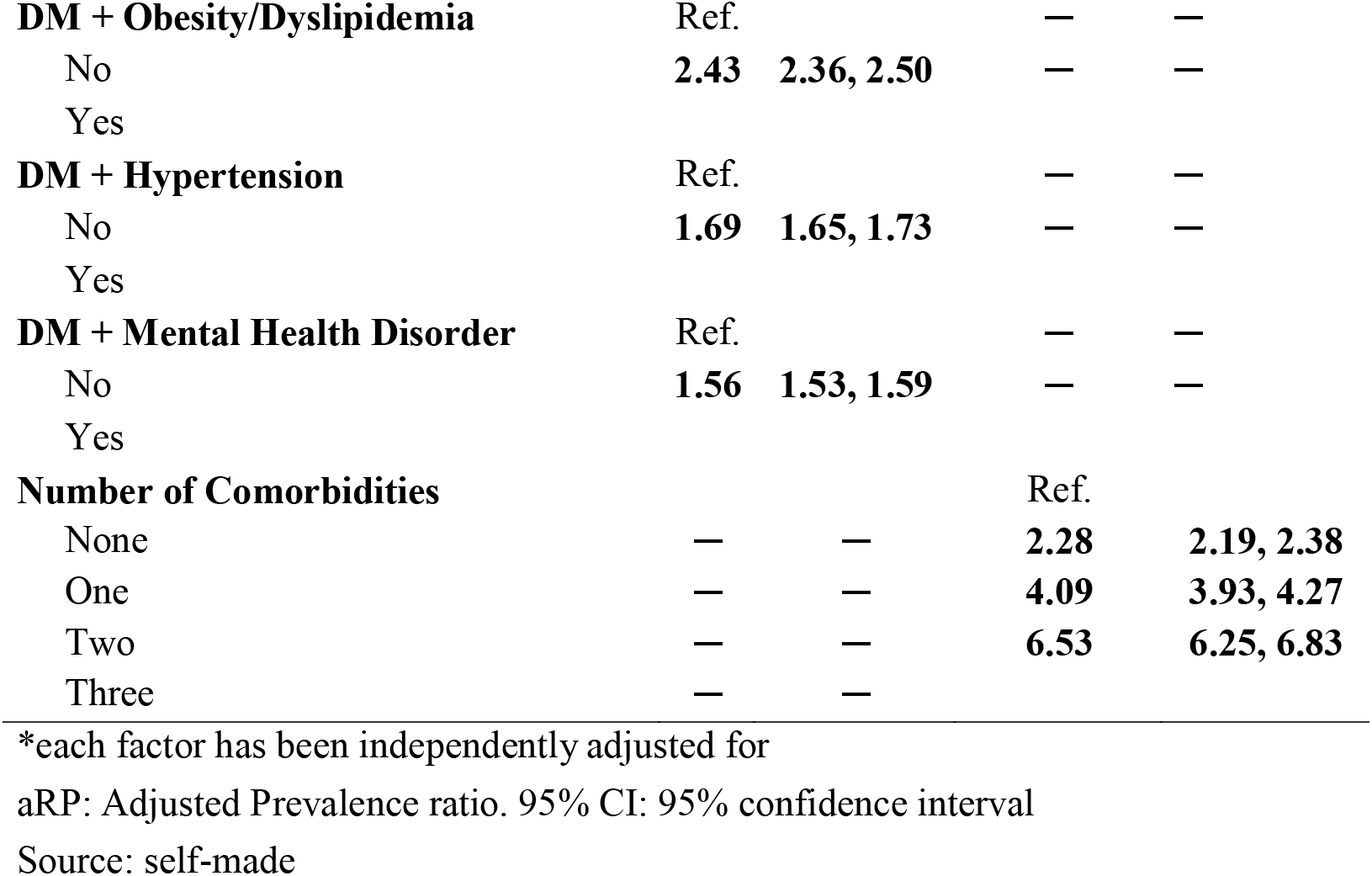
Factors Associated with the Frequency of Medical Consultations in Patients with Diabetes Mellitus: Regression Models 1 and 2.

## Discussion

### Sex and Number of Consultations

This study showed that men with DM are more likely to receive more medical consultations in the last quarter than women. This finding could be explained by several reasons. Firstly, men may experience a higher incidence of diabetes-related complications, requiring more medical consultations. Previous studies have suggested that men with diabetes may have a higher prevalence of comorbidities, such as cardiovascular and renal diseases, which could increase the need for frequent medical care ^(9,10)^. Another possible explanation is that men might have more time or availability to attend medical appointments, especially if they have fewer family or work responsibilities that interfere with their ability to seek medical care ^(11)^.

It is also important to consider that cultural and social factors may influence these differences. In some contexts, men may be more willing to report severe symptoms or insist on receiving medical care. At the same time, women may prioritize the needs of other family members over their own, resulting in fewer medical visits for themselves ^(12)^.

### Age and Medical Consultations

The study showed that the probability of receiving three or more medical consultations in the last quarter increases with age, peaking in the 60-69 age group, with a slight decrease in this probability for patients aged 70 or older. Several explanations for this trend are possible. Firstly, patients aged 60-69 are often at a critical stage in the progression of diabetes and its complications. In this age group, diabetes-related comorbidities such as cardiovascular diseases, neuropathy, and nephropathy tend to manifest with greater severity, requiring more frequent medical consultations for management. These patients may be more active and willing to seek medical care to maintain their quality of life and prevent severe complications^(13,14)^.

In contrast, the slight decrease in the probability of receiving more consultations among patients aged 70 or older could be due to several reasons. As people age, they may face greater physical and logistical barriers to accessing healthcare services, such as mobility issues, lack of adequate transportation, and physical limitations that make frequent medical visits difficult. Additionally, older patients may have different priorities and approaches to healthcare. Some may prefer less intensive and more palliative approaches focused on symptom management and quality of life rather than continuous medical monitoring. It is also possible that some older patients perceive a lower need for frequent medical care, especially if they believe that medical interventions will not significantly improve their quality of life ^(15– 17)^.

### Region and Medical Consultations

Patients residing outside Metropolitan Lima are more likely to receive three or more medical consultations in the last quarter, with this probability being exceptionally high in the Jungle region. This finding may be due to several interrelated factors influencing access to and demand for healthcare services in the different areas of the country.

Firstly, the higher probability of receiving more consultations in the Jungle may be related to the epidemiological and demographic characteristics of the region. In rural and remote areas such as the Jungle, the prevalence of infectious diseases and other chronic conditions may be higher, increasing the need for frequent medical consultations. Additionally, the population in these regions may have more comorbidities that require more intensive follow-up and management ^(18)^. Similarly, specialized services outside Lima may be limited, leading patients to receive more consultations at local health centers to compensate for the lack of specialist access ^(19)^.

Socioeconomic conditions may also play a role. Patients in regions like the Jungle may have fewer economic resources to access private services or pay for expensive treatments, leading them to rely more on the public health system and thus have more visits to government-funded health centers ^(7)^.

### Time Since Diagnosis and Number of Consultations

The analysis revealed that as the time since diabetes diagnosis increases, the probability of receiving three or more medical consultations in the last quarter decreases. This finding may seem counterintuitive, but several possible explanations and factors could contribute to this trend.

Firstly, patients diagnosed more recently may be in a phase of adjustment and stabilization of their condition. During the first few years after diagnosis, it is common for patients to require more frequent follow-up to adjust treatment, monitor response to medications, and receive education on disease management. This initial stabilization phase generally involves more medical visits to ensure blood glucose levels are adequately controlled and to prevent early complications ^(20)^. In contrast, patients with a long-standing diagnosis may have achieved a greater degree of stability in managing their diabetes. These patients likely have established more effective self-care routines and have greater knowledge and experience in managing their condition ^(21)^.

Additionally, patients diagnosed longer ago may have developed a stronger relationship with their healthcare team, allowing for more efficient disease management through less frequent but more effective consultations. These patients may also adhere more to their treatments and follow medical recommendations better, reducing the need for frequent medical interventions ^(22)^.

It is also important to consider the impact of patient fatigue. Patients with many years of diabetes management may experience “treatment fatigue,” where the motivation to attend frequent medical appointments decreases over time. This may lead to fewer medical visits as patients become more comfortable managing their disease independently and seek less regular medical support ^(23)^.

### Type of Diabetes and Number of Consultations

The study also found that patients with T2DM are less likely to receive three or more medical consultations in the last quarter than those with T1DM. This finding can be attributed to several reasons.

Firstly, the nature and management of T1DM and T2DM differ. T1DM is an autoimmune condition where the pancreas produces little or no insulin, requiring intensive and constant management with exogenous insulin. Patients with T1DM often need more frequent follow-ups to adjust insulin doses, monitor blood glucose levels, and manage health fluctuations. This complex and constant management increases the need for regular medical visits ^(24)^. In contrast, T2DM generally develops more slowly and is associated with lifestyle factors and insulin resistance. While T2DM also requires continuous management, patients can often control their condition with lifestyle modifications, oral medications, and, in some cases, insulin. The management of T2DM can be less intensive once blood glucose levels stabilize, resulting in fewer frequent medical visits than T1DM ^(25)^. Patients with T1DM and T2DM may also have different perceptions and health-seeking behaviors. Due to the severity and unpredictable nature of their condition, patients with T1DM may be more inclined to seek regular medical care ^(26)^ than those with T2DM.

Another possible explanation is the difference in associated complications. Patients with T1DM are at higher risk of acute complications, such as diabetic ketoacidosis, which require immediate and frequent medical attention. While patients with T2DM also face risks of complications, these tend to be more chronic and progressive, such as cardiovascular diseases and neuropathy, which may not require the same level of frequent medical visits once long-term management plans are established ^(27)^.

Finally, diabetes management programs and support available may differ between types of diabetes. Healthcare systems may have more structured and intensive support programs for patients with T1DM due to the critical need for close control and continuous monitoring. This may not be as pronounced for patients with T2DM, who may receive a more general approach to managing their condition ^(28)^.

### Comorbidities and Number of Consultations

Patients diagnosed with hypertension, mental health problems, and obesity are more likely to receive three or more medical consultations in the last quarter. This trend is especially pronounced in patients with obesity. Several possible explanations for these findings exist.

Hypertension is a common comorbidity in patients with diabetes and can significantly complicate disease management. The presence of hypertension requires stricter and more frequent monitoring to prevent complications such as strokes, kidney failure, and heart diseases. Patients with hypertension often need regular adjustments in their medication and close follow-up to control both blood pressure and glucose levels, increasing the frequency of medical visits ^(29)^.

Mental health disorders, such as depression and anxiety, are common in patients with chronic diseases like diabetes. These problems can complicate diabetes management due to decreased adherence to treatment, lack of motivation for self-care, and reduced ability to follow a rigorous health regimen. Patients with mental health problems may need additional support, including frequent medical consultations to manage their psychological conditions and diabetes. Moreover, mental health problems can increase the perception of symptom severity, leading to greater utilization of healthcare services ^(30,31)^.

Obesity and/or dyslipidemia are significant risk factors for numerous diabetic complications and other chronic diseases. Obese patients often have higher insulin resistance, making blood glucose control more difficult. Additionally, obesity is associated with a higher risk of developing complications such as cardiovascular diseases, sleep apnea, joint diseases, and mobility problems. These complications require intensive and multidisciplinary medical follow-up, increasing the frequency of medical consultations ^(32)^.

It is important to highlight that these comorbidities often do not present in isolation. The simultaneous presence of hypertension, mental health problems, and obesity can have a synergistic effect, where the interaction of these conditions exacerbates the severity and complexity of diabetes management. This combination can result in an even greater need for monitoring and treatment adjustment, significantly increasing the frequency of medical consultations ^(33)^. Additionally, each additional comorbidity increases the patient’s clinical management complexity. Patients with multiple comorbidities, such as hypertension, obesity, and mental health problems, require more intense and coordinated care to manage each of these conditions adequately. This includes frequent medication adjustments, continuous symptom and complication monitoring, and regular visits to assess health status and treatment efficacy ^(6,34)^.

Furthermore, comorbidities often require different treatments that can interact with each other. Managing these pharmacological and therapeutic interactions requires careful and frequent monitoring by healthcare professionals to avoid adverse effects and optimize treatment outcomes. This need for constant adjustment and evaluation of treatment interactions can increase the frequency of medical visits ^(35,36)^.

### Contribution to the Field

This study significantly public public health by identifying factors how many medical consultations patients with mellitus received. Using a big, varied database from SIS in Peru, it offers detailed understanding into how different demographic, clinical and geographic traits influence healthcare services. These results provide crucial information for planning and optimizing public health resources.

Additionally, the study highlights the importance to considering comorbidity when managing diabetic patients. The proof that those with more than one comorbidity visit doctors more often highlights the necessity for broad and coordinated care approaches. Such findings can guide design public health programs tackling both mellitus and other associated illnesses, boosting medical care efficiency and effectiveness. Identifying obesity as a particularly burden emphasizes the urgent importance for targeted weight control interventions in managing diabetic patients effectively.

The results from this study underscore the importance to adapting healthcare services to tackle regional disparities in access and usage. Places like the Jungle have more frequent medical visits, pointing to specific challenges faced by patients there. To craft effective public health policies for Peru’s varied populations, policymakers must consider these regional inequalities when forming strategies aimed at diminishing social factors that affecting care quality and system efficiency equitably across all regions. This involves providing a fair supply of accessible and efficient services, regardless of geographic a patient seeks care. Such policies guarantee optimal support throughout the continuum of care.

### Limitations of the Study

Despite this study’s significant contributions, several limitations should be considered when interpreting the results. Firstly, the study’s cross-sectional nature prevents establishing causal relationships between the evaluated variables. Another important limitation is the potential lack of information on socioeconomic and behavioral factors that may affect the use of healthcare services, as variables such as income level, access to transportation, social support, and health-seeking behavior could significantly impact the frequency of medical visits.

## Conclusions

This study has identified several key factors influencing the frequency of medical consultations received by DM patients affiliated with SIS in Peru. The findings show that male sex, age, residence outside Metropolitan Lima, and comorbidities are significantly associated with more medical consultations.

Adapting healthcare services to address regional disparities in access to and use of medical services is crucial. Public health policies should focus on individuals with specific characteristics or those in certain areas requiring greater care. Improving healthcare infrastructure and ensuring the availability of specialized services in these areas could reduce barriers to access and improve the quality of care.

## Data Availability

The data supporting the findings of this study can be accessed by the original research paper at the follow link: https://www.datosabiertos.gob.pe/dataset/afiliados-activos-en-el-seguro-integral-de-salud-con-diagn%C3%B3stico-de-diabetes-mellitus-sis

## Acknowledgments

A special thanks to the members of Tropical Diseases Research Institute, Universidad Nacional Toribio Rodríguez de Mendoza de Amazonas (UNTRM), Amazonas, Peru for their support and contributions throughout the completion of this research.

## Financial Disclosure

This study is self-financed.

## Conflict of interest

The authors declare no conflict of interest.

## Informed consent

It was not necessary to obtain informed consent in this Study

## Authors’ contribution

**Víctor Juan Vera-Ponce:** Conceptualization, Investigation, Methodology, Software, Data Curation, Formal analysis, Writing - Original Draft, Writing - Review & Editing

**Joan A. Loayza-Castro**: Investigation, Resources, Project administration, Writing - Original Draft, Writing - Review & Editing

**Rafael Tapia-Limonchi:** Validation, Visualization, Funding acquisition, Writing - Original Draft, Writing - Review & Editing

**Enrique Vigil-Ventura:** Supervision, Methodology, Writing - Review & Editing

## Notes

### Competing Interest Statement

The authors have declared no competing interest.

